# Rapid evaluation of neutralizing antibodies in COVID-19 patients

**DOI:** 10.1101/2020.09.01.20185447

**Authors:** Pingping Zhang, Baisheng Li, Wei Min, Xiaohui Wang, Zhencui Li, Yong Zhao, Huan Zhang, Min Jiang, Huanying Zheng, Chao Yang, Wei Zhang, Le Zuo, Qi Gao, Zhengrong Yang, Yanzhao Li, Tiejian Feng, Changqing Lin, Qinghua Hu, Tie Song, Ruifu Yang

## Abstract

The ongoing coronavirus disease 2019 (COVID-19) pandemic calls for a method to rapidly and conveniently evaluate neutralizing antibody (NAb) activity in patients. Here, an up-conversion phosphor technology-based point-of-care testing (UPT-POCT) and a microneutralization assay were employed to detect total antibodies against the receptor-binding domain of severe acute respiratory syndrome coronavirus 2 (SARS-CoV-2) spike protein and NAb activity in COVID-19 patients’ sera, respectively, in order to determine if UPT-POCT could be used as a surrogate method for rapid evaluation of serum NAb activity in COVID-19 patients. In total, 519 serum samples from 213 recovered and 99 polymerase chain reaction re-positive (RP) COVID-19 patients were used in this report. We found that UPT-POCT reporting values correlated highly with NAb titers from 1:4 to 1:1024, with a correlation coefficient *r* = 0.9654 (*P* < 0.001), as well as protection rate against RP (r = 0.9886, P < 0.0001). As a significant point for reducing re-positive rate, UPT-POCT values of 4.380 ± 2.677, corresponding to NAb titer of 1:64, may be appropriate as an indicator for evaluating high efficiency of protection. This study demonstrates that the quantitative lateral flow based UPT-POCT, could be used to rapidly evaluate NAb titer, which is of importance for assessing vaccine immunization efficacy, herd immunity, and screening patient plasma for high NAbs.

---

The on-going coronavirus disease 2019 (COVID-19) pandemic continues to cause global political and societal upheaval, echoing the impact of historical plague outbreaks.^1^ For the prevention and control of this black swan event, the best strategy is effective vaccination.^2^ However, from experience of previous coronavirus pandemics, the development of an effective vaccine still faces some unpredictable obstacles. Herd immunity implement is highly controversial,3 and it has been proven that in nonhuman primates, previous infection with severe acute respiratory syndrome coronavirus 2 (SARS-CoV-2) can protect against re-challenge.^4^ Also, some infected patients treated with convalescent plasma therapy have shown improvement, making it a potential therapeutic option.^5^ However, to determine whether antibodies produced either during herd immunity, vaccination, or in convalescent plasma are protective, a rapid surrogate method to evaluate the protective effect of antibodies against SARS-CoV-2, instead of traditional neutralization tests, is urgently needed. Here, we employed up-conversion phosphor technology-based point-of-care testing (UPT-POCT) to quantitatively detect total antibodies, and a microneutralization assay to evaluate the neutralizing activity of the antibodies from both recovered and polymerase chain reaction (PCR) re-positive (RP) COVID-19 patients. The results demonstrate that UPT-POCT could be used as a surrogate method for quickly evaluating neutralizing antibody (NAb) activity in COVID-19 patients, for potential vaccination efficacy evaluation, and a tool for implementing immunity passports.^6^

## Methods

All patients were diagnosed, treated, and discharged according to Guidelines for Diagnosis and Treatment for Novel Coronavirus Pneumonia (Seventh Edition) issued by the National Health Commission of the People’s Republic of China.^7^ Serum samples were collected from recovered patients, and serial sera were collected from all PCR RP and some recovered patients. In total, 519 serum samples from 213 recovered (98 males and 115 females) and 99 RP (58 males and 41 females) COVID-19 patients in Guangdong province of China were collected. The Median age of patients was 40 years (range from ten months to 86). All sera were analyzed by quantitative immunochromatographic strip and microneutralization assay to determine total antibodies and antibody neutralizing activity, respectively. This research was approved by the ethics committees of Shenzhen Center for Disease Control and Prevention (QS2020060007).

The UPT-POCT kit used in this study, Novel Coronavirus 2019-nCoV Antibody Test (Up-converting Phosphor Immunochromatographic Technology), was jointly developed by our lab and Beijing Hotgen Biotech Co., Ltd., and has been licensed by the National Medical Products Administration (NMPA) of China. In this kit, the receptor binding domain (RBD) and S1 protein expressed in eukaryotic cells were employed, respectively, as the test strip-coating antigen and up-conversion phosphor nanoparticle (UCP)-labeling protein. This kit was developed for quantitative detection of total antibodies in COVID-19 patients. For the UPT-POCT assay, a 100 μL mixture of 10 μL serum and 90 μL diluting buffer were applied to the sample window on the strip. After 15 min of lateral flow, the result was read by a specific biosensor for test and control bands, with results reported as the ratio of test (T) to control (C) signals (T/C ratio).^8^ The microneutralization assay was performed according to previous reports (**see supplementary material**).^9,10^

## Results

All 519 serum samples were first used to verify if the antibody levels detected by UPT-POCT correlated with neutralizing titers (**Figure 1 and supplementary Table S1**). As shown in Figure 1, T/C ratios correlated well with neutralizing titers from 1:4 to 1:1024, with a correlation coefficient *r* = 0.9654 (*P* < 0.001). Most (61.3%, 318/519) of the serum samples had neutralizing titers between 1:32 and 1:128, with T/C ratios between 2.735 ± 1.860 and 5.019 ± 2.941. The positive correlation observed between T/C ratios and neutralizing titers indicate that T/C ratio values may be used as a surrogate marker for rapidly evaluating antibody neutralizing activity. Previous studies have confirmed that angiotensin-converting enzyme 2 (ACE2) is the main receptor recognized by the RBD region of the spike protein of SARS-CoV-2.^11^ UPT-POCT employs the RBD fragment as the coating protein on the testing strip and S1 protein as the UCP-labeling antigen, targeting antibodies with potential neutralizing activity; thus T/C ratio values could be a surrogate biomarker for evaluating an antibody’s neutralization potential. Next, we postulated whether NAbs in RP patients have lower protective effect relatively to those in non-RP (NRP) recovered patients. To assess this, we analyzed the correlation between T/C ratios, RP rates, and accumulated RP (ARP) rates at each neutralizing titer (**Figure 2 and supplementary Table S2-S3**). Here, the ratio of the number of RP cases to the sum of RP and NRP patients at each condition was defined as the RP rate; while the ARP rate at a specific neutralizing titer is defined as the ratio of APR patients to the sum of ARP and accumulated NRP patients at titers above the designated titer [ARP rate = ARP/(ARP + accumulated NRP)]. As we collected serial sera from RP and some NRP patients with more than one neutralization result, the lowest neutralizing titer and corresponding T/C ratio were chosen for further analysis. In total, 312 cases, including 99 RP and 213 NRP patients, were analyzed. As shown in **Figure 2A and Table S2**, at T/C ratios equal to or greater than 4.380 ± 2.677 (neutralizing titers ≥ 1:64), RP rates at each neutralizing titer steadily decreased and remained below 30%, with half of all cases (46%, 144/312) meeting those criteria. The correlation between T/C ratios and ARP rates is illustrated in **Figure 2B and Table S3**. As with the RP rates, at T/C ratios equal to or greater than 4.380 ± 2.677 (neutralizing titers ≥ 1:64), ARP rates showed a constant decline from 24% (34/144) to 0%, covering almost half (46%, 144/312) of all cases. For the remaining patients (54%, 168/312), with neutralizing titers below 1:64, ARP rates were approximately 30%.

**Figure 1.**
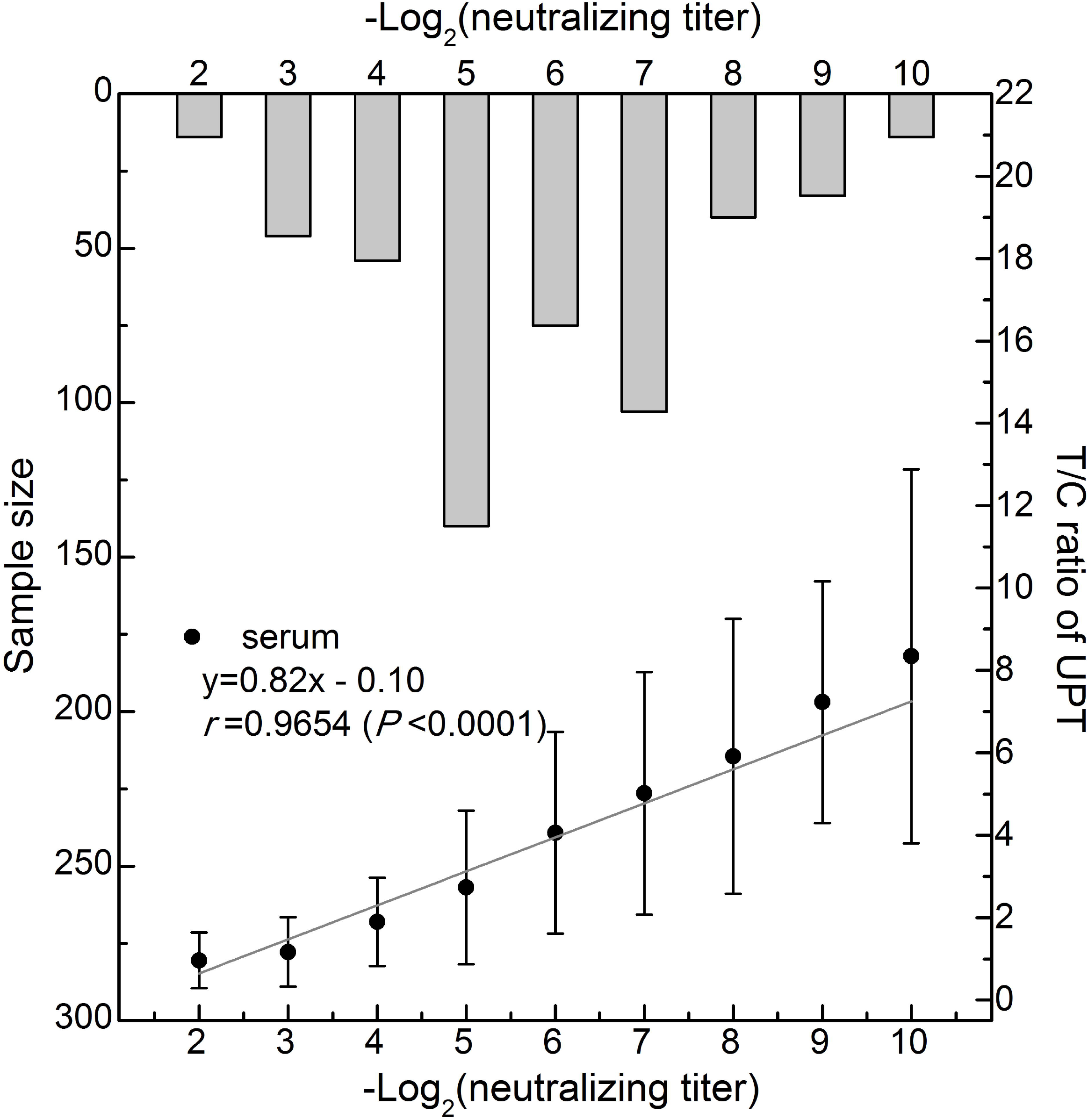
Correlation between T/C ratios and neutralizing titers. X-axis indicates neutralizing titers in negative log^2^. Left Y-axis shows the corresponding sample sizes (light grey bars), right Y-axis represents the T/C ratios reported by UPT-POCT.

**Figure 2.**
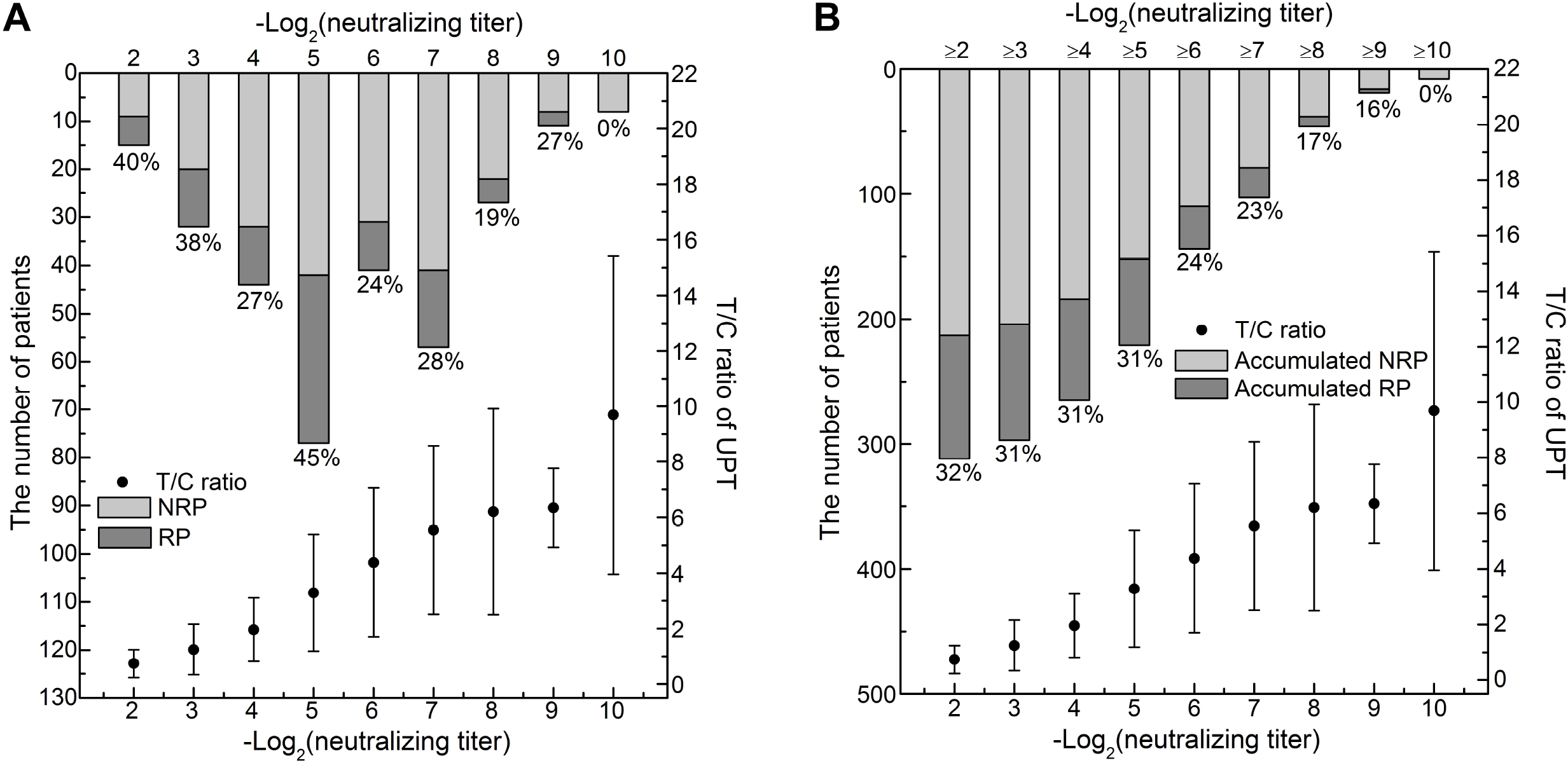
Correlation between T/C ratios and re-positive rates. Plot of the correlation between T/C ratios and RP rates (A) and accumulated RP (ARP) rates (B). X-axis indicates neutralizing titers in negative log^2^. Left Y-axis shows the number of NRP (light grey bars) and RP patients (dark grey bars), or accumulated values. Right Y-axis represents T/C ratios reported by UPT-POCT at a specific neutralizing titer. Percentage figures in A and B indicate RP and ARP rates, respectively, calculated as the ratio of the number of RP cases (or accumulated cases) to the sum of RP and NRP patients (or accumulated cases).

As shown above, RP rates with T/C ratios from 0.738 ± 0.509 to 3.287 ± 2.102 (titers from 1:4 to 1:32) (39%, 65/168) were significantly greater than those from 4.380 ± 2.677 to 9.689 ± 5.730 (titers from 1:64 to 1:1024) (24%, 34/144). These results are consistent with the consensus that antibodies can protect recovered patients against re-positive.

Finally, the correlation between T/C ratios and protection rates against RP was explored (**Figure 3 and Table S4**) in order to evaluate whether UPT-POCT could be used as a convenient surrogate biomarker to rapidly assess the neutralizing activities of antibodies in COVID-19 patients and in human populations actively vaccinated or passively via herd immunity. Here, we define the protection rate against RP at each neutralizing titer as the ratio of the total number of cases less APR patients to the total number of cases [(total cases - ARP patients)/total cases]. As shown in Figure 3A, protection rates reached 89% and 78% at T/C ratios equal to or greater than 4.380 ± 2.677 (corresponding titers ≥ 1:64) and 3.287 ± 2.102 (corresponding titers ≥ 1:32), covering approximately half (46%, 144/312) and three quarters (71%, 221/312) of total cases, respectively. Significantly, for T/C ratios ≥ 0.738 ± 0.509 (corresponding titers ≥ 1:4), protection rates exceeded 68%, demonstrating that the antibodies can protect recovered patients against re-positive to a significant degree as long as they can be detected. Linear regression analysis between T/C ratios and protection rates against RP were performed, resulting in a correlation coefficient *r* = 0.9886 with T/C ratios from 0.738 ± 0.509 to 9.689 ± 5.730 (corresponding to the neutralizing titers from 1:4 to 1:1024) and protection rates against RP from 68% to 100%. This reveals that the quantitative UPT assay can give a value for predicting levels of protection rates against RP, which could be employed as a convenient surrogate method to the microneutralization assay for the quick evaluation of the neutralizing activity of antibodies in target serum.

**Figure 3.**
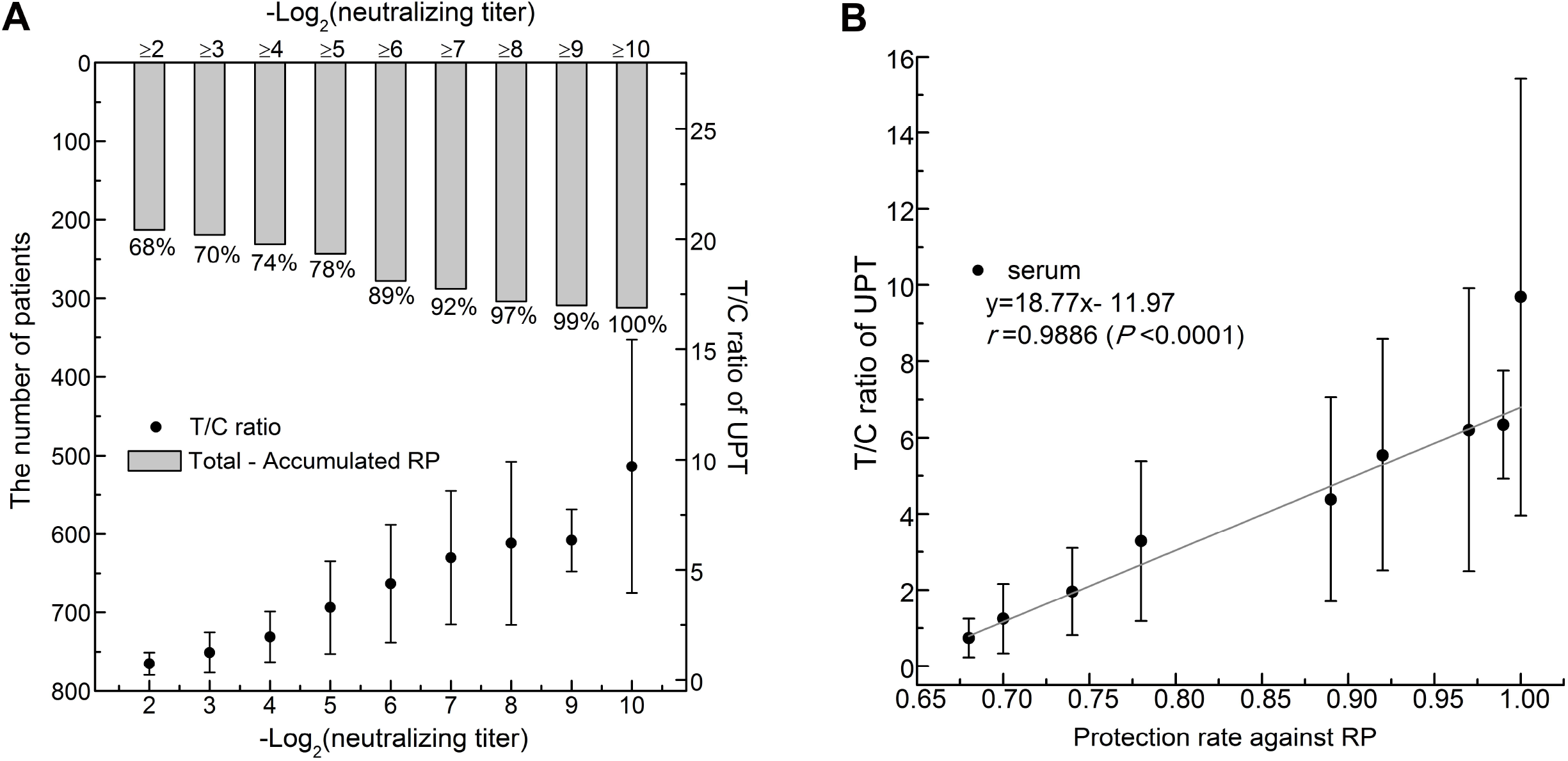
Correlation between T/C ratios and protection rates. (A) Protection rates at each neutralizing titer. X-axis indicates titers in negative log^2^. Left Y-axis shows the total number of patients less accumulated RP patients at a specific neutralizing titer (grey bars). Percentages are the ratios of the number of cases at that titer to the total number of cases, which is defined as the protection rate against RP. Right Y-axis shows T/C ratios of the UPT assay. (B) Linear regression between T/C ratios and protection rates against RP.

## Discussion

Although monoclonal antibodies with neutralizing activity have been isolated from convalescent COVID-19 patients,^12^ clinical improvement by convalescent plasma therapy is controversial,^5,13^ partly because of the lack of criterion for the measurement of neutralizing titers due to its complex operation.^9^ The rational design of effective vaccines for COVID-19 requires identification of neutralizing antibodies that protect against SARS-CoV-2 after vaccination,^2^ a process that would be facilitated by a simple immunoassay that reflects virus neutralization. By analyzing 519 serum samples from 213 recovered and 99 PCR RP COVID-19 patients in this study, T/C ratios of UPT-POCT were found to correlate highly with SARS-COV-2 neutralizing antibody titers (*r* = 0.9654, *P* < 0.001), proving a promising biomarker for quick evaluation of an antibody’s neutralization potential. The neutralizing titer for 61.3% of the sera were between 1:32 and 1:128, with T/C ratios between 2.735 ± 1.860 and 5.019 ± 2.941, which is consistent with previous reports on the relatively low neutralizing titers of convalescent plasma,^14,15^ demonstrating the risk of recovered patients becoming RP cases.

Neutralizing titers correlated with protective effect^2^ with a linear relationship to T/C ratios, thereby demonstrating the possibility of utilizing UPT-POCT to assess protective effect for immunity. For the 312 recovered or PCR RP COVID-19 patients, T/C ratios for neutralizing antibody titers showed high correlation with protection rates against RP. A T/C ratio of 4.380 ± 2.677 (neutralizing titer = 1:64) was the threshold for reducing RP rates, and is therefore recommended as an indicator for evaluation of protective effect for vaccination. Furthermore, UPT-POCT can give a more accurate value for protection rates due to the consistent linear relationship, which is, to some extent, superior to the more time-consuming and semi-quantitative microtneutralization assay. Targets of protective NAbs are primarily found on the spike protein as it is the sole viral surface protein required for entry into the host cell, and there is great interest in NAbs that block the binding between RBD and ACE2.^16,17^ However, NAbs that do not bind to the RBD have also been found^18,19^ and epitopes in the spike occupied by non-NAbs^20^ may have an influence on the neutralizing titer of serum samples. Therefore, although RBD-specific IgG titers have been found to be partly correlated with SARS-CoV-2 neutralizing titers (*r* = 0.622),^13^ the significance of this correlation cannot be determined due to the possible influence of non-NAbs or other types of antibodies, such as IgA and IgM.^21^ In this study, we detected total antibodies against RBD using a double antigen sandwich method and the proteins S1 and RBD, showing that T/C ratios correlated with neutralizing titers and protection rates against RP. It can be inferred that NAbs against RBD are primarily responsible for protective effect and according to the recent studies, anti-RBD titers are recommended as a surrogate method to identify suitable plasma donors based on analysis of convalescent plasma and asymptomatic patients’ plasma.^22^ These facts will aid the development of more simple methods for evaluating neutralization, which could be helpful for preventive and control measures against COVID-19, such as the rational design of effective vaccines, convalescent plasma therapy screening, and evaluation of protective passive herd immunity in a population. As immunity against SARS-CoV-2 is believed to be short-term and proliferation of the virus can occur at any time of year, the current pandemic has been predicted to last for a number of years.^23^ To relieve the profoundly negative impact of the pandemic on economic recovery and social stability, immunity passports have been suggested to enable some individuals protected against secondary infection to return to work. However, this strategy will be difficult to implement due to the lack of information regarding the relationship between neutralizing antibodies and subsequent infection.^6^ Our study has found that results of point-of-care testing and titers of neutralization tests can be valuable references in monitoring protection against re-positive, and could be valuable tools for the implementation of immunity passports.

## Data Availability

The data used to support the findings of this study are available from the corresponding author upon request.

## Author contributions

RY conceived the study. RY, TS, QH and CL designed the experiments. BL, WM, XW, PZ, ZL, YZ, HZ, MJ, HYZ, WZ, LZ, QG, ZY, YL, and TF performed the experiments. PZ, RY and CY analyzed the data. PZ and RY wrote the manuscript.

## Acknowledgments

This study was supported by Beijing Municipal Science & Technology Program (Grant No. Z201100005420022), The National Key Research and Development Program of China (Grant No. 2018YFC1200502), National Natural Science Foundation of China (82041030), Sanming Project of Medicine in Shenzhen (No. SZSM201811071), The China National Science and Technology Major Projects Foundation (No. 2017ZX10303406), and Shenzhen Key Medical Discipline Construction Fund (No. SZXK064).

